# An Empowered Transfer Learning Model for Predictive Classification of Lung Cancer

**DOI:** 10.1101/2025.05.21.25328060

**Authors:** Syed Thouheed Ahmed, Satheesha Tumakur Yoga, Lakshmi Hassan Nagaraja, Sandeep Kumar Mathivanan, R. Sangeetha, Saurav Mallik

## Abstract

Lung cancer detection and treatment is processed using upgraded medical tools and radiology experts from multiple medical data sources. The challenge in decision making is dependent on experts understanding on a given electronic health records or datasets. In this paper, we have proposed an improvised approach of lung cancer classification based on intensity driven RoI selection from the Lung Images Database Consortium Image Collection (LIDC-IDRI), Cancer Imaging Archive (CIA) datasets. The technique is developed on label customization and annotating the vulnerable RoI regions. The approach is optimized for higher dimensionality mapping of RoIs. The technique deploys a feedback-based upgrading and monitoring approach via transfer learning framework. The trained dataset from RoI optimizer is updated to customized learning models for decision transfer and decision-making capabilities. The proposed technique is deployed on CoVNET framework and has demonstrated the accuracy of 97.84% under a 60:40 training testing-based learning model.

## I. INTRODUCTION

Cancer, an uncontrolled growth on the organ (internal or external) due to the mutation is the major concern in public health. The origin and spread of cancer is unfortunately uncontrolled by the prevision medicines. According to World Health Organization (WHO) a record of 10 million deaths are reported in 2020 with 1.8 million deaths due to lung cancer as second most cancer worldwide with 2.8 million deaths due to breast cancer in first place. The lung cancer occurrence is due to major concerns such as smoking, radiation exposure and genetic history. The medical field has advanced in providing drugs and medical solutions in treating the cancer. The technical contribution via modern day technological enhancements has assured an ease in the diagnosis and decision support. The techniques such as basic image processing using pattern recognition [1] is considered as the initial advancements to provide diagnosis and cancer lesion extraction. with time, the dataset of lung cancer and the repositories have evolved under the centralized remote processing environment supporting technologies such as machine learning and deep learning with neural networks as the primary input phase for diagnosis [2].

Given the severity of lung cancer and the need for research using modern techniques, the proposed manuscript is aimed to provide a reliable solution for CT lung-cancer dataset classification using transfer learning model. The objective of transfer learning [3] is to provide a self-evolving machine learning model with improvising with each implementation cycle. The Convolutional Neural Networking (CNN) model is added to support the diagnosis and classification of the lung cancer via transfer learning. The approach provides a classification of lung cancer into three major categories such as normal, benign and malignant as shown in Fig. 1. The predominant dataset representation is recorded in Magnetic Image Resonance (MRI), Computed Tomography (CT) and X-ray towards technical research, whereas in this research, the CT datasets are considered on the primary computation. The lung cancer identification and the clinical solutions [4] is provided based on the promising approaches of computer-aided diagnosis (CAD) for analyzing the CT images in an effective manner.

**FIGURE. 1:**
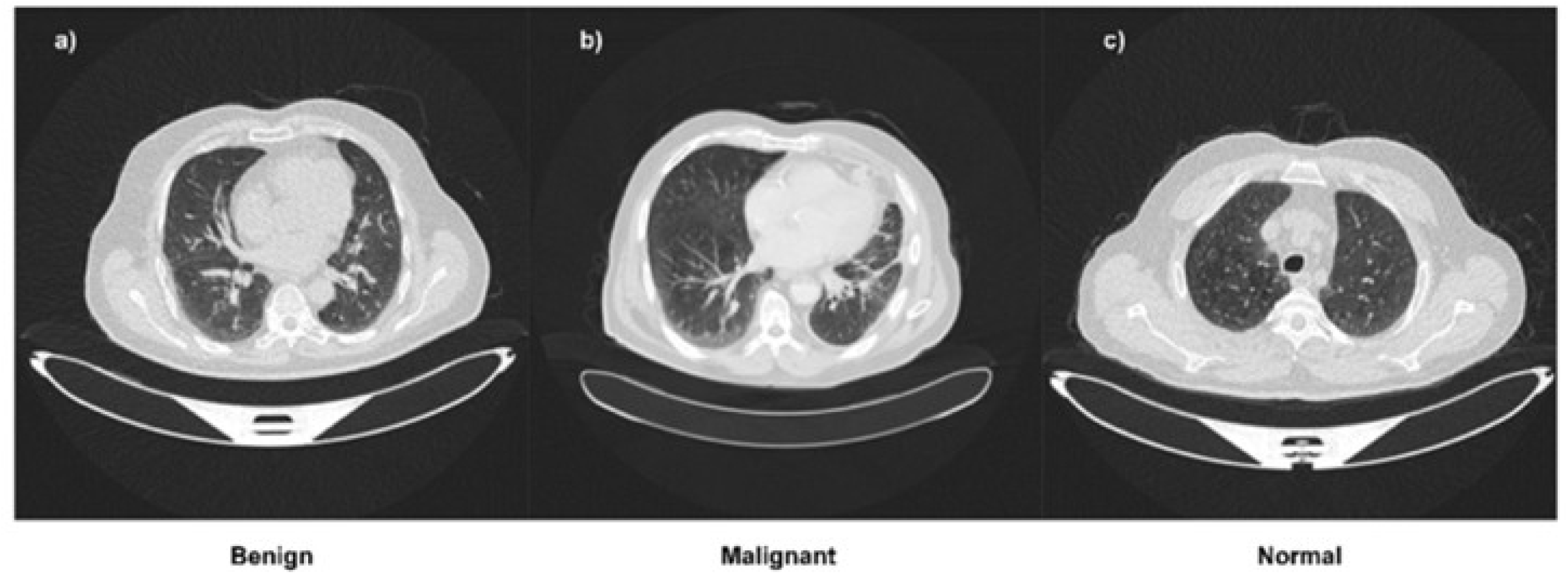
CT Lung image with Benign, Malignant and Normal representation.

The machine learning and deep learning models associated with the lung cancer based lesion classification, segmentation and prediction is based on the static dataset training and hence requires a computational complexity on the implementation. The deep learning and machine learning models widely use CT as a reliable and simple dataset for processing. The CT datasets include features and attributes such as annotations cum segmentations with nodules, tumor (lesion) and other abnormalities highlighted. The publicly available datasets are Lung Nodule Analysis (LUNA16) datasets [18] and the Cancer Image Archive (CIA) are most commonly used datasets for the computation.

The manuscript is designed with an objective to provide a reliable and effective solution for lung cancer classification specific to CT image dataset. The proposed framework is supported by a modified CNN layer represented as Convolutional Network (CoVNET) for improvised classification. The technique is supported by a transfer learning model via feedback-based approach for collective improvisation of learning model with each implementation cycle. The approach has specified the intensity driven RoI extraction of CT images datasets and processed in CoVNET model for effective classification and decision-making capabilities over the labelling of RoI patterns. The technique is novel in design towards the customization of the processing CT datasets via feedback-based learning model.

The manuscript is organized with the domain introduction and contribution in section I followed by section II with a detailed and recent developments in literature and case studies on lung cancer classification via CT datasets. Section III includes the proposed methodology and the proposed model design in section VI with a detailed representation of transfer learning model and the modified CNN layers. The results and outcome is discussed in section V with a detailed discussion on the CoVNET based transfer learning model comparative analysis. The manuscript is summarized and concluded in section VI followed by references.

## II. LITERATURE SURVEY

Lung Cancer the second most prominent cause of death world by WHO and a detailed study [5] is a testimony from International Association for the Study of Lung Cancer staging project. The technological enhancement in machine learning and deep learning approach. The reviews and detailed literature of various screening techniques in CT dataset for lung cancer detection is presented in [6] via deep learning, [7] via clinical datasets and [8] via open datasets for classification. The deep learning approaches such as Lung-EffNet [9] and U-Net based segmentation [10] for effective classification and dataset optimization towards developing the modified AlexNet-SVM classification model at 97.98%. The approaches assures the behavioral validation cum segmentation of the CT images into the multiple slices for effective processing and training of the model.

An effective lung carcinoma based classification is discussed in [11] using CNN and CAD based derived datasets. The carcinoma datasets are higher in resultant and positive prediction ratio compared to the regular three classes of categorization and the similar clinical support on dataset review is reported in [12] on addressing the studies using machine learning for lung cancer classification and diagnosis. Thus categorizing the cancer (lung) into malignant or benign is a challenging task and a technique in [13] has demonstrated a machine learning based approach for optimizing the datasets and classifying it accordingly using Cancer Cell Detection using Hybrid Neural Networking (CCDC-HNN). With the inclusion of Artificial Intelligence (AI), the lung-cancer based immunotherapy is proposed in [14] with modern advancement in validating the learning models, whereas the in [15] the TVFx-based COVID-19 diagnosis on the lungs provide a classification using feature thresholding technique.

The X-Ray based multi-class lung cancer classification is validated in deep learning approach in [16] considered as chest X-ray (CXR). The dataset plays a vital role and hence the combinations of data representation using MRI, CT and X-Ray under CSV and comma-separated files. In [17] a data structural representation is validated on clustering the image datasets for effective identification of dissimilar groups. The studies reported in this survey are aligned with the scope of proposed technique with respect to image segmentation and rebuilding a machine learning model via transfer learning.

The approaches in [19] and [20] has demonstrated the COVID-19 disease classification using chest X-rays and Bayesian Deep Learning (BDL) model. The approach is validated on DenseNet-121 and EfficientNet-B6 with 70,000+ images on training and validation data-sample size.

The major set-back in the overall techniques is using a single-source or single repository dataset for processing, training and validation of the model, whereas the change in dataset has resulted in changing the overall decision support of the learning model. The collective representation in this manuscript has addressed the challenge of synchronizing third-party datasets from open repositories and the pre-processed dataset for decision support via transfer learning

## III. METHODOLOGY

The proposed methodology is developed on the data labeling and customizing third-party CT datasets for processing. The archives from the public CT images dataset is acquired under the common server/repository based on local computational unit. The local servers provide a primary labeling on the CT lung images in classifying the datasets into normal, malignant and benign as shown in Fig. 1. The dataset labelling further assures the dataset attributes consistency for customizing the formulated labels into a segmented index of parameters and ratio values. The typical consideration on the label is to associate the training dataset with a classifier and categorize into the three categories. The third party datasets from external sources are further included in re-aligning the attributes labeled and creating a patch annotation and classified optimizations on derived datasets as demonstrated in Fig. 2. The selection of customized labels and Region of Interest (RoI) is extracted for the given datasets towards convolutional neural networking (CNN) models computation. The CNN model is demonstrated and remodified as shown in Fig. 3. The process of input images (CT) based computation under CNN and decision support assures the models reliability in securing higher classification ratio of the lung cancer into the normal, malignant and benign. The process is further supported by feedback based learning approach to improvise the patch annotation and labeling on the CT images. The outcome of CNN decision layer is further justified with a public domain computational unit of neural network operated by third party cum open network channel for model backup and recovery. The detailed model design is demonstrated in successive section.

**FIGURE. 2:**
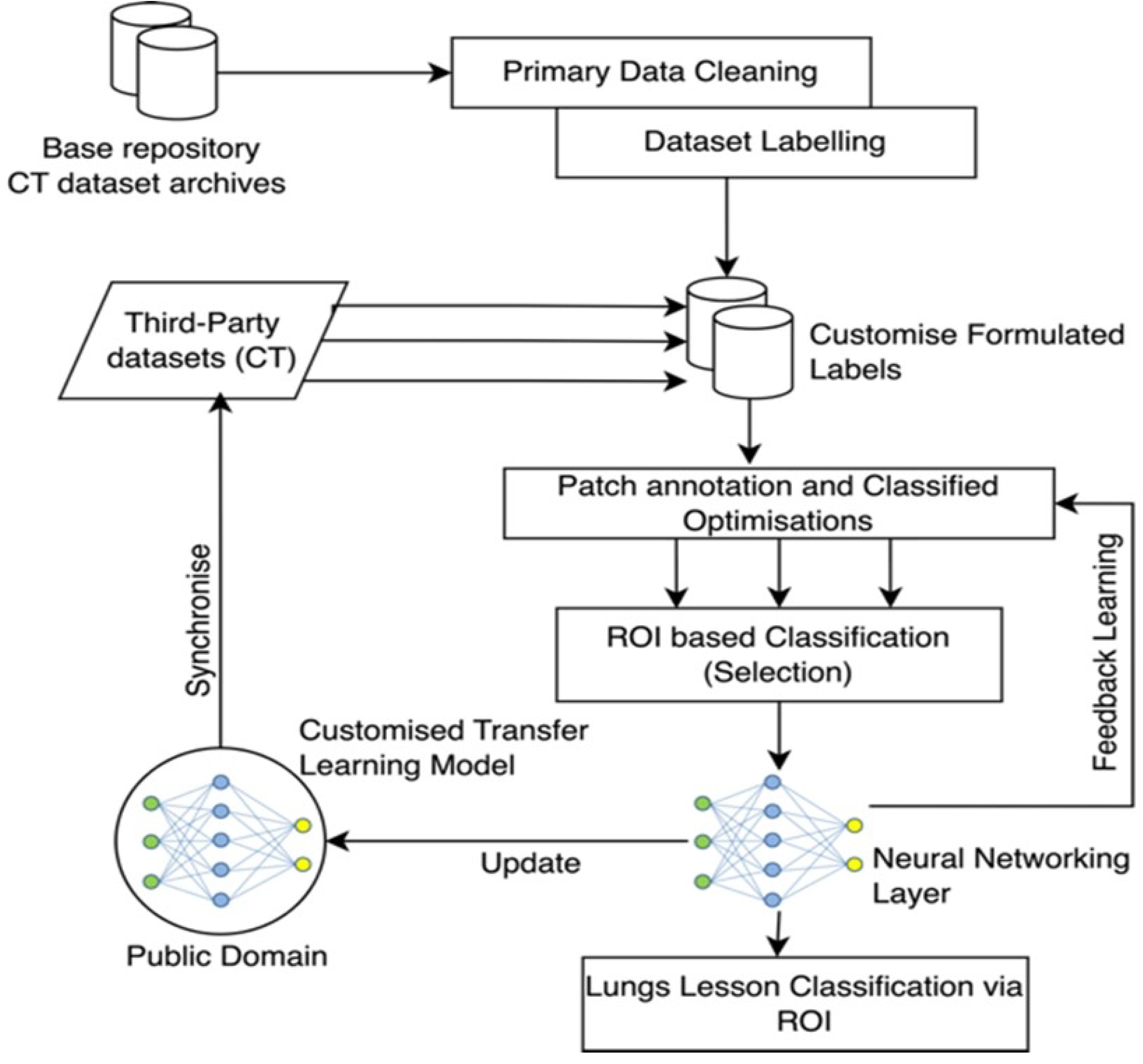
Proposed System Architecture.

**FIGURE. 3:**
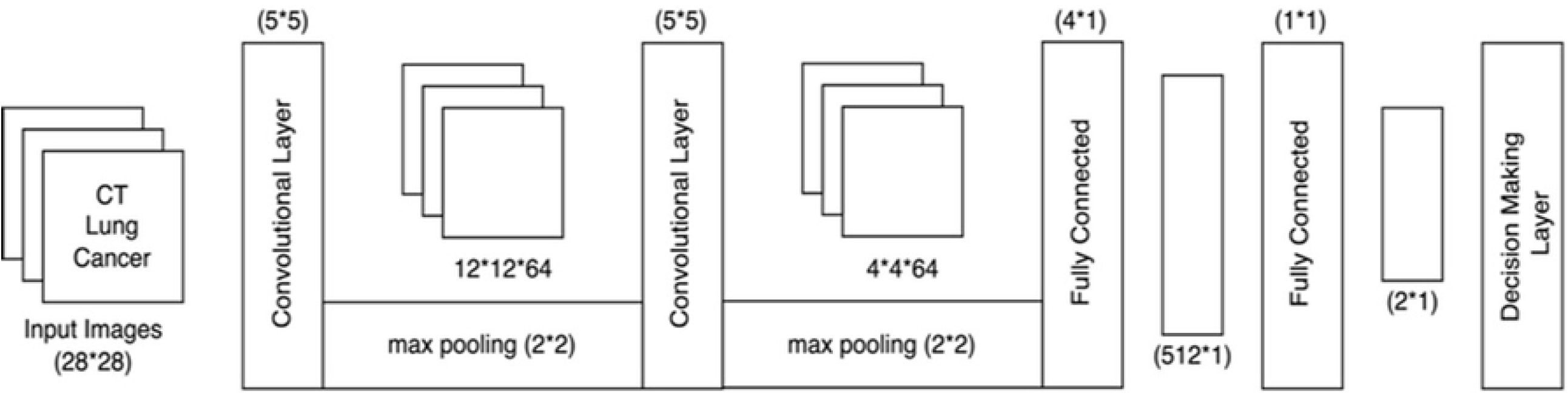
CNN Model of proposed system.

## IV. MODEL DESIGN

### A. PROBLEM STATEMENT

The lung cancer dataset based decision making is dependent on experts’ analysis and manual intervention. The process accompanies multiple challenges such as dataset availability, expert availability and demand of patient follow-up with particular medial expert. The major research challenge is to fetch single information point for lung cancer medical dataset classification and decision making. Consider the given dataset (*D*_*X*_) in the orientation to customize multiple information as (*I*_1_, *I*_2_, *I*_3_….*I*_*n*_) such that (*D*_*X*_ ⇒ *I*_*i*_ / *i* ≠ 0) at a given instance of time (*t*). The value of each information set is projected with corresponding values of segments (*S*_*X*_) as (∀*S*_*X*_ ≅ [*D*_*X*_ U *I*_*i*_]) at each computational time of dataset. Thus to expand the correspondence, the dataset (*D*_*X*_) needs to be classified for independent information/feature representation.

In the scenario of classification (*C*), under regular operations, the relative classification matrix (*C*_*M*_) is dependent on the type of datasets (*D*_*X*_) and order of feature selection (*F*_*S*_) such that 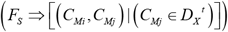 where each passing interval of information is correlated to the customized dataset attribute (*D*_*X*_) bound to time (*t*). Hence the relevance of each informative stream is left unexposed for decision support. A dedicated solution towards classification is required for lung cancer datasets. The solution in this manuscript is based on intensity driven RoI of dataset and relevant feature extraction.

### B. INTENSITY DRIVEN ROI EXTRACTION

In this paper, the intensity driven RoI based feature extraction is proposed. The proposed framework is influenced via dataset RoI segmentation and customization using CNN framework as represented in Fig. 2. The input stream of cancer dataset is processed as follows

#### CASE STUDY

Consider the use case of dataset prepressing and customization. Consider the input stream of CT datasets from Cancer Image Archive (CIA) repository and represented as (*D*_*X*_), where (*D*_*X*_) is the universal and large visible dataset. The processing and cleaned dataset is represented as (*D*_*K*_) is extracted as (*D*_*K*_ ⊆ *D*_*X*_). Thus on generalized representation the large dataset orientation can be customized as shown in Eq. 1.

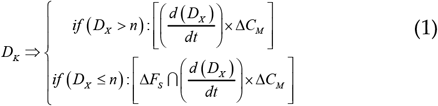

Where according to Eq. 1, the relevance of (*D*_*X*_) is extracted based on (*D*_*X*_) and (*n*), such that (*n*) is the value of dataset request for primary processing. Thus from the understanding of (*n*) influences the (*D*_*K*_) is computed. Typically, the extracted dataset (*D*_*X*_) is further recommended for customization of formal label (*L*). The labels array such that the feature values and matrix representation is explored as shown in Eq. 2.

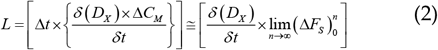

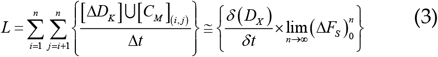

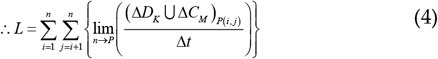

Thus according to Eq. 2, the formal labels (*L*) are bound on secondary parameters of extracted datasets (*D*_*K*_) with reference to classification matrix (*C*_*M*_) as defined by the processing agent. In general, the hypervisor (*C*_*M*_) values at given time (*t*) is correlated to universal dataset (*D*_*X*_) based feature selection (*F*_*S*_). The process of corelationship mapping represents in Eq. 2 and Eq. 3 respectively, where (∀ *D*_*K*_≅ [*D*_*X*_ U *F*_*S*_]) on extensive compression of values. The optimization of labels are further represented in Eq. 3 and Eq. 4, where (lim(*n → P*)) is introduced. (*P*) is the progression threshold value set by the training vector to extract maximum intensity of RoI from dataset (*D*_*K*_). The inference of (*P*) on (*D*_*K*_) and (*C*_*M*_) is bound with time (*t*) such that (∀*P →*[*D*_*K*_ ∩*F*_*S*_]) at a larger feature interference. Typically, the relationship of (*F*_*S*_ → *C*_*M*_ ⇒ ∃*D*_*K*_) and (∀*D*_*K*_ ⇒ *C*_*M* (*i, j*)_)at a bounding time (*t*) for optimized computation.

The overall inference from each (*D*_*K*_) _(*I*, *j*)_ is dependent on the structural representation of feature-set (*F*_*S*_) with respect to classification matrix (*C*_*M*_) on time (*t*) interval. The intensity of RoI(R) is extracted with interdependency matrix as shown in Eq. 5. The (*R*) dependencies are evaluated as below.

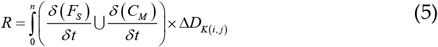

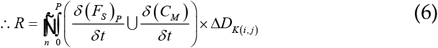

Thus the rational values of (*R*) RoI is extracted from Eq. 5 and consolidated in Eq. 6 as each (*F*_*S*_) and (*C*_*M*_) are dependent on time (*t*). The (*D*_*K* (*i, j*)_) value assures the operations in multiple-dataset consolidation and collection to optimize the RoI selection.

### C. TRANSFER LEARNING AND DECISION MAKING

The computed RoI extraction process (*R*) is streamlined and feed as input to the convolutional neural network (CNN) model. The CNN computes detailed RoI framing as shown in Fig. 3. The input (28 × 28) is layered into (5 × 5) ⊕ (5 × 5) convolution layer with intermediate max pooling of (2 × 2) with phase-1 of (12 ×12 × 64) and phase-2 of (4 × 4 × 64). The resultant values are streamlined to the fully CNN layer generating (4 ×1) and (1×1) ratio for decision support. Technically, the CNN model optimizes the operational vector of all these parameters within the ratio scale of operations.

The input vector and resultant matrix of two segmented layers are further customized and resultant in the operational behavioral analysis. The outcome analyses the decision support on customizing the input medical images via customized datasets (*D*_*K*_). The decision support here further justifies the lesion classification in lungs. The computational and transfer learning on model is extended to a public domain via customized transfer learning model (*T*_*L*_) as shown in Eq. 7.

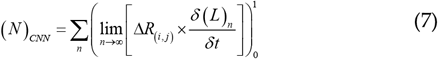

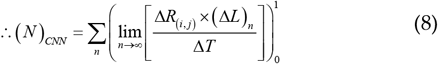

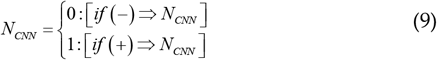

Thus according to Eq. 8 and Eq. 9, the customization is resultant to a decision support, whereas from Eq. 8, the transfer learning (*T*_*L*_) model customization is processed and evaluated. The transfer learning model (*T*_*L*_) is resultant under open domain of computation as shown in Eq. 10.

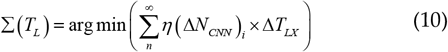

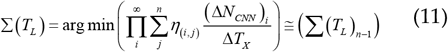

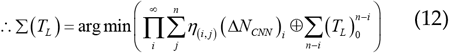

Thus from Eq. 11 and Eq. 12, the consolidation is optimized for the model of operation. The aggregation is processed from each building (Δ*N*_*CNN*_) model to collectively coordinate in developing a transfer learning model (Δ*T*_*L*_) such that (∀ *T*_*L*_ ∈ Δ *η*_(*i,j*)_) with (∃*N*_*CNN*_ ∈*T*_*L*_) at a given interval (*t*). Technically, the upgrading process of each learning model is cumulatively built and transferred to a training dataset to retain higher order of decision support.

## V. RESULTS and DISCUSSIONS

The outcome of the proposed technique is to assure the computational classification of the lung cancer based on the RoI based transfer learning. The dataset is extracted from the Lung Images Database Consortium Image Collection (LIDC-IDRI), Cancer Imaging Archive (CIA) datasets on 1025 lung cancer images (CT data format) or CT images of size 767 MB (Collectively). The experiment setup is two cluster based A100 NVIDIA 40GB SXM GPU enabled configuration for CoVNET (*T*_*L*_) based CNN model training and validation. The initial dataset is LIDC-IDRI and third party repositories are Kaggle CT lung cancer Stage-1 and Stage 2 datasets.

### A. CoVNET MODEL EVALUATION

The initial peak of training testing accuracy is demonstrated in Fig. 4 with a training at 60% of LIDC-IDRI datasets and testing with 40% under 200 epochs. Technically, the defined ratio of training and testing accuracy is relatively consistent on the training accuracy with a diffraction of moving average 2.3 at the testing phase. Whereas the individual losses at the 50, 100, 150 and 200 epochs are recorded in Fig. 5 with RoI density extraction. The improvisation ratio of training and testing losses are minimized with a difference of minimal 0.34 epochs ratio. The resultant validation is subjected with the implementation benefits of Transfer learning (*T*_*L*_) and collectively represented in Fig. 6.

**FIGURE. 4:**
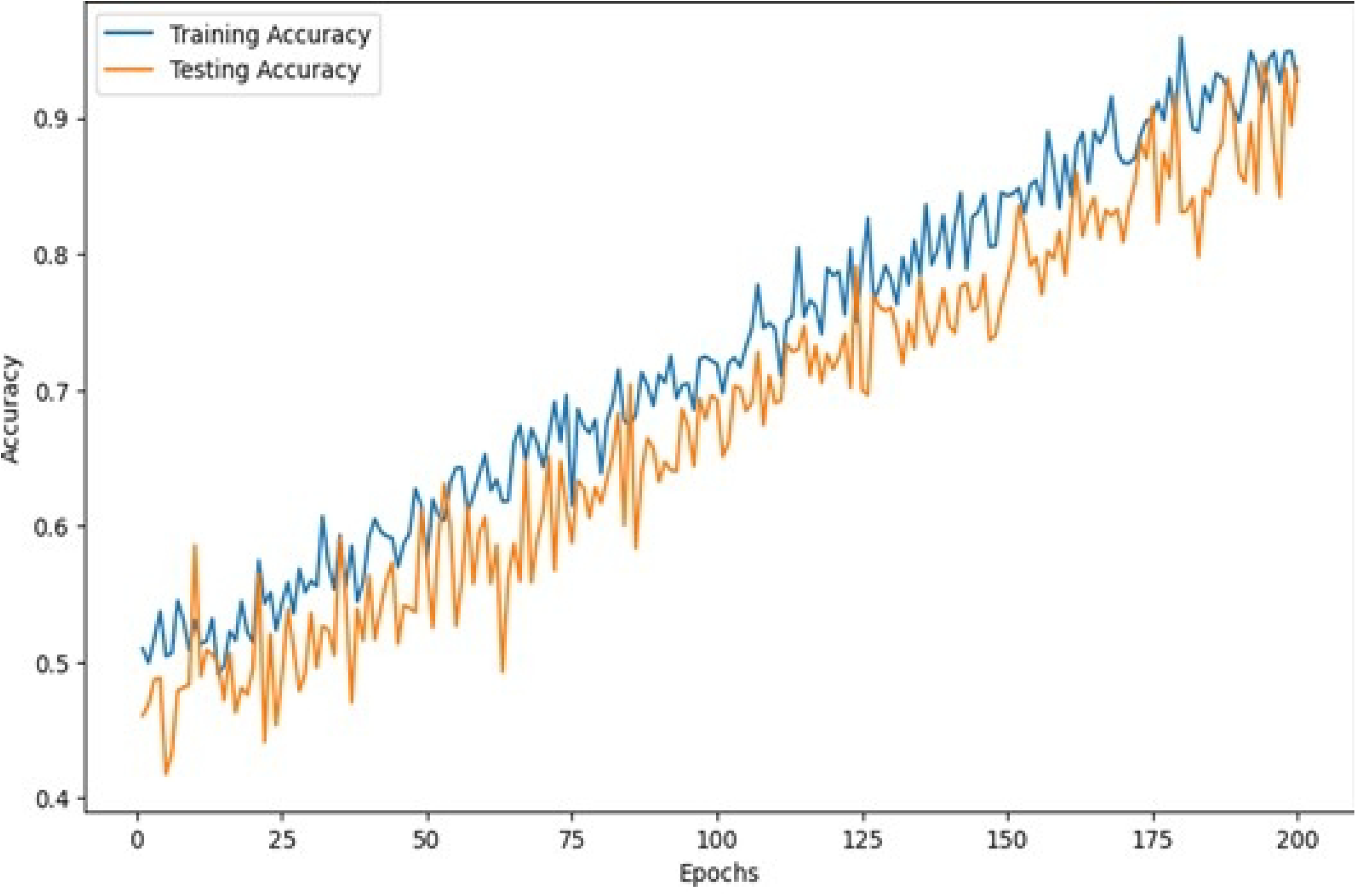
Training accuracy of the proposed CT Lung cancer image classification (Collective)

**Fig. 5:**
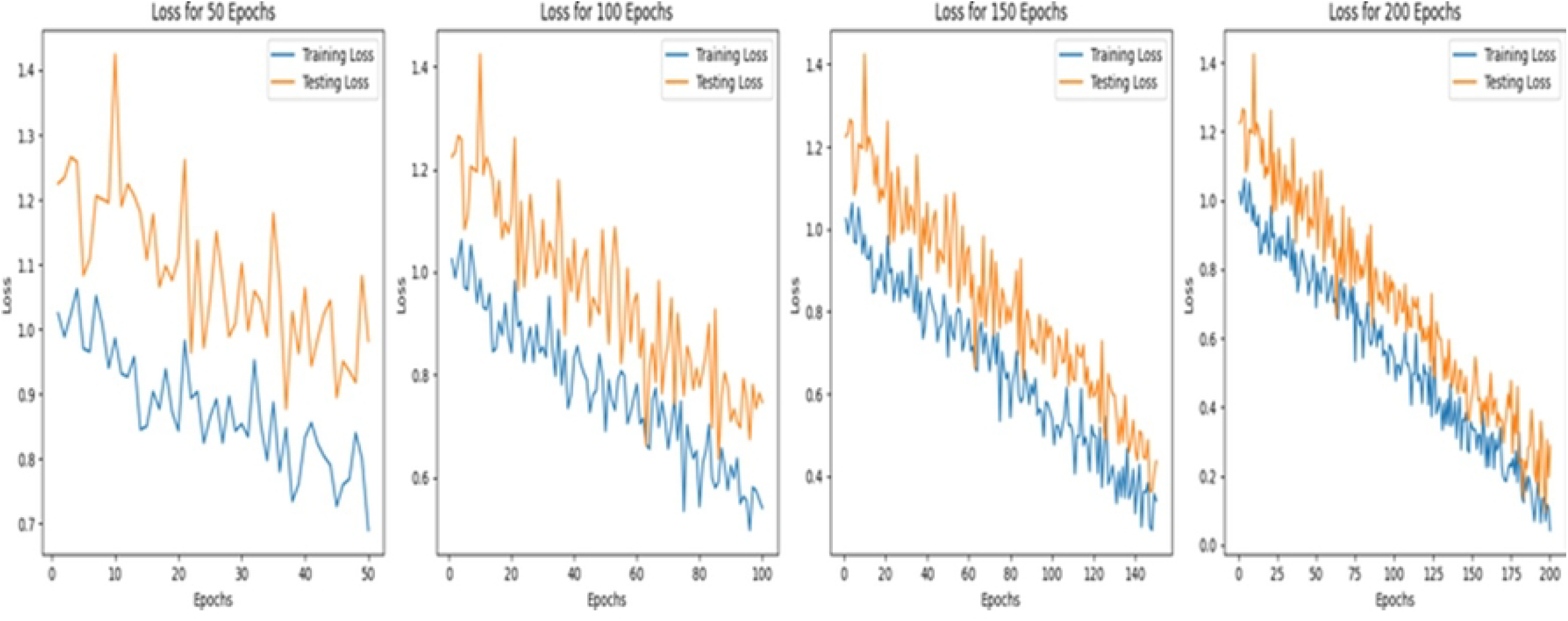
Training v/s Testing Loss of the proposed CT Lung cancer image classification with 50, 100, 150 and 200 epochs.

**Fig. 6:**
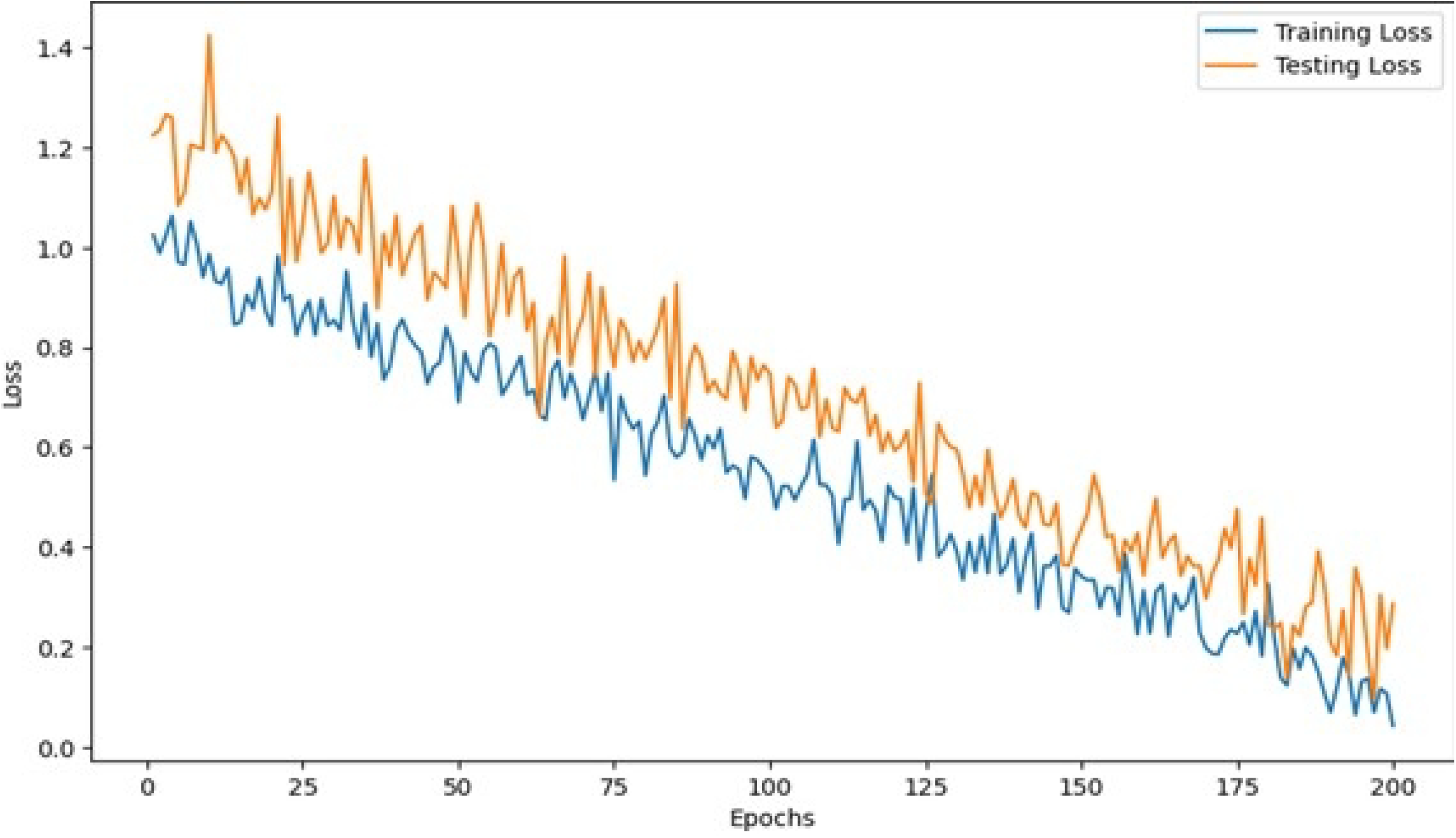
Training v/s Testing Loss of the proposed CT Lung cancer image classification (Collective)

The collective confusion matrix is evaluated on the primary classification of CT Lung cancers such as normal, malignant and benign. Thus according to the proposed transfer learning model, the representation is streamed on 30units to maximize the decision making capabilities of the proposed model. The confusion matrix is demonstrated in Fig. 7 and represents the predictive validation of each classification class. The summarization format of the collective CoVNET model derived on transfer learning (*T*_*L*_) approach is represented in Fig. 8. The validation is supported with three major splits of datasets (D_X_), a primary ratio of testing and training is 60:40 ratios, followed by 70:30 and 80:20 splits. The training ratio at 60% and 70% selection has achieved a higher decision ratio with 200 epochs whereas the three selections in testing is been validated at near saturation for the predictive classification as shown in Fig. 8. The detailed split ratio is demonstrated in Table. 1 with a classification accuracy prediction. The proposed CoVNET based transfer learning model has further justified the performance matric as shown in Table. 2. The confusion matrix in Fig. 7 is derived on the Table. 2 evaluation ratio for plotting the decision support.

**FIGURE. 7:**
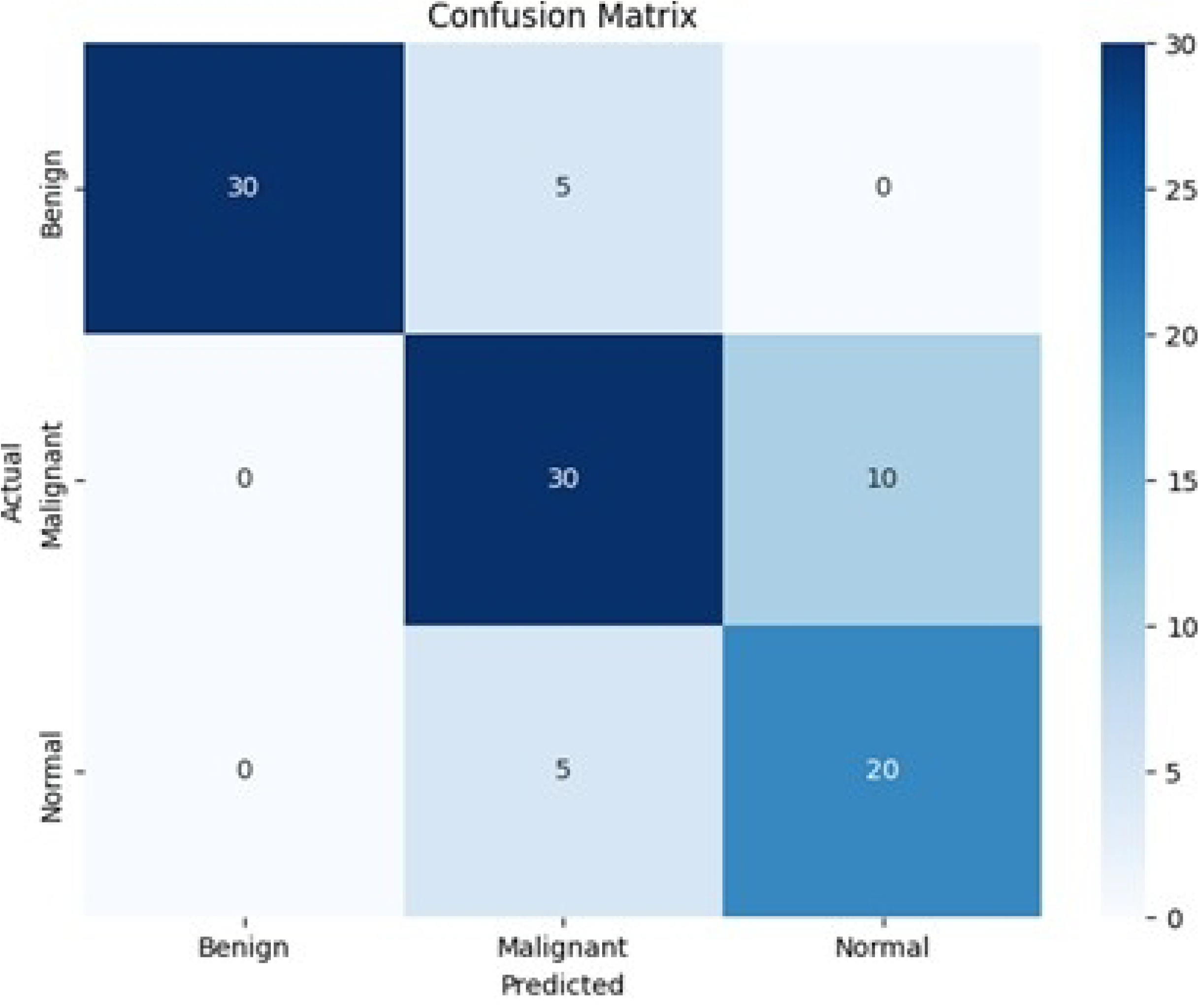
Confusion matrix of plotting a classification of lung cancer based on occurrence (i.e.) Benign, Malignant and Normal based on CoVNET proposed model.

**FIGURE. 8:**
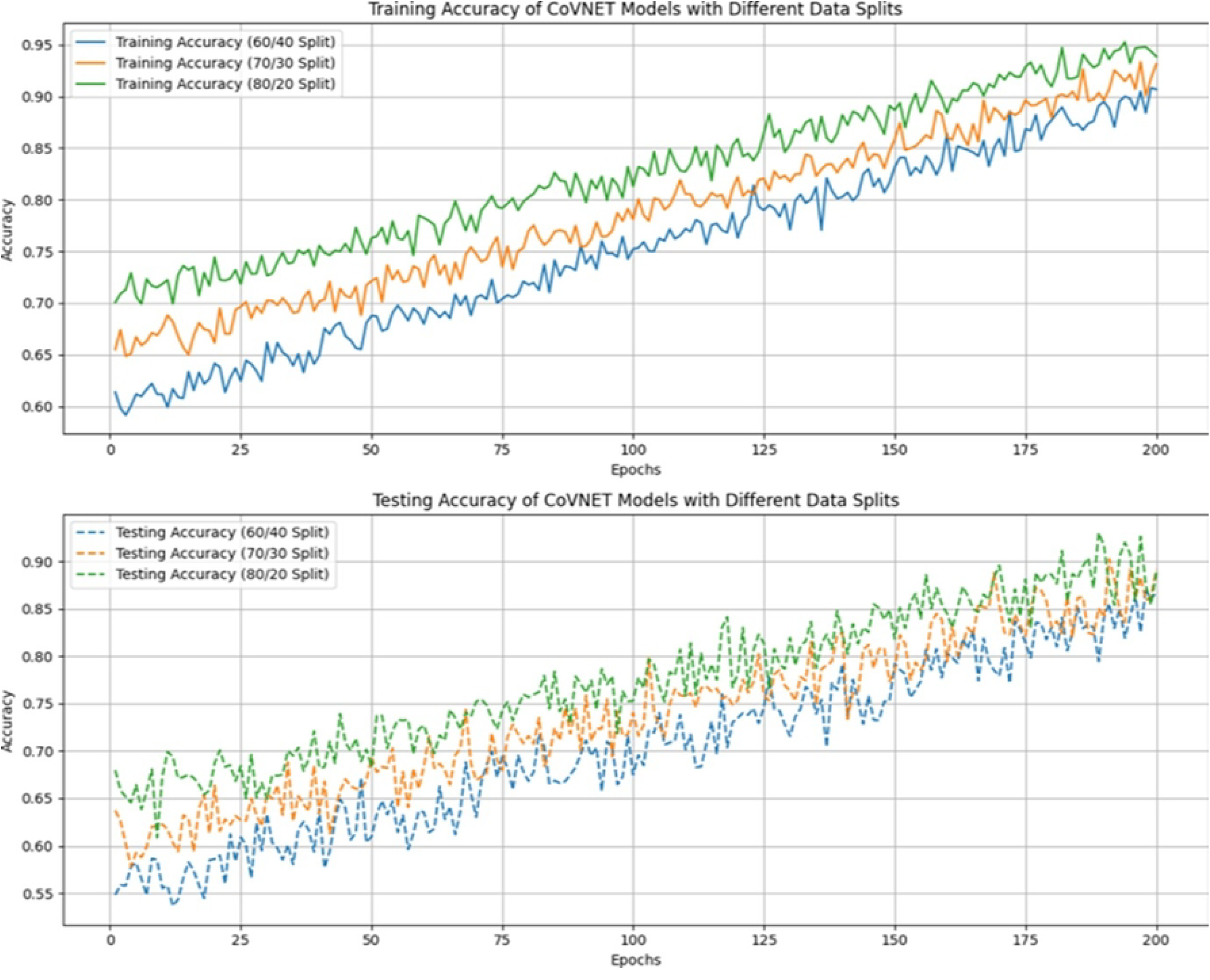
Training v/s Testing Loss.

**TABLE 1:** DATASET TRAINING AND TESTING WITH RESPECT TO THE SPLITTING RATIO AND CLASSIFICATION CLASSES TO PREDICT THE CLASSIFICATION ACCURACY.

| Class | Split | Phase | Sample Input size | Processed Input size | Classification accuracy (%) |
| --- | --- | --- | --- | --- | --- |
| Normal | 60/40 | Train | 312 | 102 | 92.23 |
| Benign |  |  | 211 | 109 | 91.97 |
| Malignant |  |  | 289 | 118 | 92.79 |
| Normal |  | Testing | 78 | 52 | 93.31 |
| Benign |  |  | 66 | 48 | 92.29 |
| Malignant |  |  | 61 | 24 | 93.03 |
| Normal | 70/30 | Train | 341 | 112 | 91.43 |
| Benign |  |  | 271 | 118 | 92.17 |
| Malignant |  |  | 189 | 105 | 91.99 |
| Normal |  | Testing | 55 | 55 | 92.11 |
| Benign |  |  | 39 | 36 | 92.24 |
| Malignant |  |  | 40 | 39 | 92.06 |
| Normal | 80/20 | Train | 391 | 212 | 93.53 |
| Benign |  |  | 227 | 128 | 92.45 |
| Malignant |  |  | 173 | 112 | 92.19 |
| Normal |  | Testing | 38 | 31 | 93.03 |
| Benign |  |  | 32 | 28 | 92.15 |
| Malignant |  |  | 29 | 22 | 93.03 |

**TABLE 2:** PERFORMANCE MATRIX OF PROPOSED COVNET TRANSFER LEARNING MODEL.

|  | Precision | Recall | F1 Score |
| --- | --- | --- | --- |
| Normal | 100 | 100 | 100 |
| Benign | 98.47 | 99.02 | 97.21 |
| Malignant | 98.21 | 98.78 | 98.32 |

### B. COMPARATIVE ANALYSIS

The proposed CoVNET based transfer learning model (Δ *T*_*L*_) is derived with the accuracy ratio of 98.78% with reference to the derivative score processed in Table. 2. The comparative analysis on the existing models is demonstrated in Table. 3. The training testing ratios at 60:40 is higher in performance ratio compared to the 70:30 and 80:20. The observation is drawn on the maximizing the transfer learning model in minimizing the computation time and maximizing the accuracy ratio. The proposed model (CoVNET-BX) is implemented on five different intervals of study with each model is interconnected to the previous instance for transfer learning the decision support accordingly. The model is further evaluated on the universal EfficientNETBX instance and GoogleLeNet with deriving the accuracy at 93.38% and 94.84% accordingly. The proposed re-modified CoVNET (collective) model is derived with an accuracy of 97.84%. The model under independent instances with is maximized on the CoVNET-B5 instance compared to the previous instances.

**TABLE 3:** COMPARATIVE ANALYSIS OF TRAINING MODEL ON (Δ*T*_*L*_) FOR ACCURACY COMPUTATION.

| Model | 60:40 |  | 70:30 |  | 80:20 |  | Dataset<br>Size<br>(MB) | Accuracy<br>(%) |
| --- | --- | --- | --- | --- | --- | --- | --- | --- |
|  | Precision | F1<br>Score | Precision | F1<br>Score | Precision | F1<br>Score |  |  |
| CoVNET-B1 | 97.11 | 98.62 | 97.47 | 98.11 | 96.32 | 97.11 | 112 | 97.28 |
| CoVNET-B2 | 97.29 | 97.19 | 96.12 | 97.74 | 96.19 | 96.89 | 178 | 97.74 |
| CoVNET-B3 | 98.31 | 99.32 | 98.18 | 97.89 | 98.11 | 97.78 | 215 | 98.62 |
| CoVNET-B4 | 98.48 | 99.34 | 98.29 | 97.11 | 98.48 | 98.12 | 328 | 98.35 |
| CoVNET-B5 | 98.47 | 99.32 | 98.27 | 97.43 | 98.21 | 98.04 | 418 | 98.63 |
| EfficientNetB0 | 97.32 | 98.11 | 97.74 | 96.12 | 98.17 | 97.12 | - | 93.67 |
| GoogleLeNet | 94.21 | 91.32 | 98.43 | 98.23 | 99.32 | 100 | 538 | 94.38 |
| **Proposed**<br>**(CoVNET)** | **98.34** | **98.84** | **97.11** | **97.87** | **97.32** | **98.43** | **767** | **97.84** |

## VI. CONCLUSION

The proposed technique (CoVNET) model is derived from the transfer learning model based on Region on Interest (RoI) extracted via the modified CNN layer. The technique has successfully derived the CT lung-cancer images classification into the predominant three classes. The Proposed CoVNET model is trained and tested on the three-split ratio of 60:40, 70:30 and 80:20 respectively. The model has provided a stable coordination of training and testing at 60:40 ratio. The overall technique has successfully derived the transfer learning model for the information building via connected ratio of CoVNET feedback towards upgrading the computational CNN model. The technique has achieved the accuracy of 97.84% in classifying the CT lung cancer. In near future, the model is further derived on multi-dimensional CT lung cancer datasets for 3D based evaluation.

## Author Contributions

All authors have read and agreed to the published version of the manuscript.

## Funding

This research is supported by University of Arizona.

## Ethics approval and consent to participate

“Not applicable”.

## Institutional Review Board Statement

“Not applicable”.

## Informed Consent Statement

“Not applicable”.

## Acknowledgements

“Not applicable”.

## Data Availability Statement

The datasets used during the current study are available from the corresponding author on reasonable request.

## Conflicts of Interest

“The authors declare no conflict of interest.”

## Notes

### Competing Interest Statement

The authors have declared no competing interest.

### Funding Statement

No funding

